# A Neo4j-Based Framework for Integrating Clinical Data with Medical Ontologies: Performance Optimization and Quality Measure Applications in Healthcare

**DOI:** 10.1101/2025.07.20.25322556

**Authors:** Sohyeon Jeon

## Abstract

**Background:** Electronic Health Records face a fundamental challenge: the semantic gap between relational data storage and clinical reasoning patterns. Traditional databases struggle with complex healthcare queries requiring multiple joins and temporal analysis, creating performance bottlenecks that limit real-time clinical applications.

**Methods:** We developed a Neo4j-based framework integrating MIMIC-IV clinical data (1,504 patients, 4,967 admissions) with SNOMED CT medical ontology through ICD-10-CM mappings. The implementation created a unified graph comprising 625,708 nodes and 2,189,093 relationships, with systematic preservation of temporal and semantic connections.

**Results:** Performance analysis demonstrated substantial improvements over PostgreSQL across five query types, with Neo4j showing 5.4x to 48.4x faster execution times. The framework successfully enabled three clinical applications: ventilator-associated pneumonia temporal analysis (revealing 47.79% pneumonia rates among ventilated ICU stays), hypertension semantic network mapping through multi-level SNOMED-CT relationships, and Medicare Part D quality measure monitoring. Notably, the system identified that 96.7% of eligible diabetic patients lacked statin prescriptions, demonstrating practical utility for healthcare quality improvement initiatives.

**Conclusion:** This graph-based approach provides a robust foundation for next-generation clinical decision support systems by bridging the gap between fragmented clinical data and integrated patient-centric analysis. The framework’s demonstrated performance advantages and practical applications in quality measure monitoring establish its potential for addressing real-world healthcare challenges while supporting the transition toward more effective, evidence-based patient care.

## Introduction

MIMIC-IV (Medical Information Mart for Intensive Care) is a publicly available dataset containing comprehensive clinical data from patients admitted to the emergency department or intensive care units (ICUs) at the Beth Israel Deaconess Medical Center between 2008 and 2019. The database includes over 65,000 ICU admissions and more than 200,000 emergency department visits, providing a rich repository for clinical research and healthcare analytics [1,2]. Adhering to HIPAA compliance through rigorous de-identification processes, MIMIC-IV enables secure data sharing within the research community, making it an invaluable resource for healthcare studies.

Meanwhile, SNOMED-CT (Systematized Nomenclature of Medicine - Clinical Terms) serves as the most comprehensive and precise clinical terminology system globally, encompassing over 300,000 concepts related to diseases, symptoms, procedures, and more. It enables semantic interoperability across electronic health records (EHRs) supporting clinical decision-making processes and facilitates the integration of diverse healthcare data sources and improves the consistency of clinical information [3,8,9]. SNOMED CT is one of a suite of designated standards for use in U.S. Federal Government systems for the electronic exchange of clinical health information and is also a required standard in interoperability specifications of the U.S. Healthcare Information Technology Standards Pane [32,33]. Studies have shown that SNOMED-CT enables semantic interoperability between healthcare systems and supports the exchange of clinically validated health data between providers [28,29]. Implementation studies across healthcare organizations have demonstrated significant operational benefits, including reduced medical errors and improved patient safety through standardized clinical documentation [29,30]. A comprehensive literature review of SNOMED-CT implementations between 2013-2020 confirmed its effectiveness in supporting clinical decision-making and improving healthcare service efficiency [31]. These evidence-based advantages have contributed to SNOMED-CT becoming a foundational terminology system for EMR implementation in U.S. healthcare organizations.

Traditional relational databases, while effective for structured data management, struggle with representing patient-specific clinical trajectories and addressing complex healthcare relationships, as integrating diverse patient data sources in a meaningful way presents significant technical limitations [11]. Their rigid structure and reliance on join operations create performance bottlenecks and limit their ability to capture complex temporal and semantic relationships inherent in clinical data. These limitations underscore the need for advanced data models that can holistically represent individual patient journeys while maintaining connections to broader biomedical knowledge [4].

To address these limitations in traditional databases, alternative approaches using modern technology have emerged. Graph database such as Neo4j offer an alternative approach by directly linking nodes through edges, enabling efficient traversal of complex relationships and scalable querying of large datasets. Unlike traditional systems, Neo4j’s graph-native architecture enables direct relationship traversal without join operations, making it particularly suitable for representing hierarchical and temporal relationships between clinical events [27]. These capabilities make it particularly suitable for integrating diverse healthcare datasets like MIMIC-IV and SNOMED-CT into a unified framework that supports patient-specific analysis [4,5,6,7].

Healthcare delivery is increasingly shifting from provider-centric to patient-centric models, requiring a more comprehensive understanding of individual patient journeys [13,14,15]. This shift highlights the need for frameworks that can integrate diverse data types – including medical records, genetic information, lifestyle factors, and real-time health data-into a holistic view of patient health. Such frameworks align with the growing demand for personalized medicine approaches by enabling multidimensional analyses of patient trajectories.

While numerous biomedical knowledge graphs such as BioCypher [16], SPOKE [17], PrimeKG [18], AlzKB [19] have significantly advanced biomedical research by elucidating molecular mechanisms and disease pathways, they primarily focus on molecular-level or disease-centric relationships. These frameworks lack integration of patient-specific clinical data, leaving a critical gap in modeling individual patient trajectories within broader biomedical contexts. Addressing this gap requires developing integrated frameworks that can effectively model patient-level clinical information while maintaining connections to existing molecular and disease knowledge.

Therefore, this study aims to develop a unified graph database that addresses three key challenges in clinical data analysis: (1) efficient representation of complex clinical relationships, (2) semantic integration of medical terminologies, and (3) scalable querying of patient trajectories. By transforming MIMIC-IV into a graph model and connecting it with SNOMED-CT concepts through standardized mappings, this research provides a foundation for advanced clinical decision support systems and personalized medicine approaches.

This study introduces a novel graph database framework that integrates MIMIC-IV clinical data with SNOMED-CT’s standardized medical terminology using Neo4j. By preserving temporal relationships between clinical events and leveraging SNOMED-CT’s semantic structure, this framework enables comprehensive analysis of both individual patient trajectories and population-level clinical pathways. Unlike previous efforts that focused on isolated aspects of healthcare data integration [10,20,21,22,23,24] or clinical standardization, this approach bridges fragmented datasets into a unified graph model capable of supporting advanced clinical decision-making systems.

## Methods

### Data Resources

MIMIC-IV version 2.2 was obtained through PhysioNet after completing the required CITI training course for accessing the database. The complete dataset contains over 65,000 ICU admissions from Beth Israel Deaconess Medical Center. For this initial study, we utilized a subset consisting of 1,504 patients with 4,967 admissions and 2,104 ICU stays, along with 12,456 Clinical Event nodes representing diagnoses, procedures, and measurements. The database is organized into three primary modules: hosp (hospital-wide data such as patient demographics, diagnoses, and procedures), icu (intensive care unit-specific data), and note. Integrating the hosp and icu modules resulted in 3,876 MAPS_TO relationships linking MIMIC-IV diagnoses to SNOMED-CT concepts.

SNOMED CT International Edition (July 2024) was used as the primary clinical terminology system, containing over 300,000 clinical concepts structured across diseases, symptoms, procedures, and other medical terms. The data is provided in RF2 (Release Format 2) format with three core components: Concepts, Descriptions, and Relationships files. We specifically utilized the Snapshot release type, which provides a complete view of all active components at the release time, rather than the Full or Delta releases. This version includes ICD-10-CM reference sets mapping over 126,000 SNOMED CT source concepts to ICD-10-CM targets. While SNOMED CT contains over 300,000 concepts, our implementation utilized a subset of 8,743 Concept nodes, 15,234 Description nodes, and established 22,567 Relationship edges between concepts, as these were the concepts that directly mapped to diagnoses and clinical events present in our MIMIC-IV patient dataset through ICD-10-CM mappings.

### Data Loading Process

The MIMIC-IV and SNOMED CT datasets were loaded separately due to their inherently different data structures, requiring distinct integration approaches within our data model. For MIMIC-IV, we implemented a hierarchical loading process that preserves the complex relationships within clinical data. The process begins with loading core patient data which contains demographic information. Admission records are connected to their respective patients using HAS_ADMISSION relationships, while ICU stays are linked to admissions via HAS_ICU_STAY relationships. Finally, clinical events such as diagnoses, procedures, and chart events are integrated while preserving their temporal relationships within the patient timeline.

SNOMED CT data integration utilized Neo4j’s native LOAD CSV functionality to directly import three essential RF2 files. These include the Concepts file containing foundational clinical entities, the Descriptions file providing human-readable terms, and the Relationships file defining semantic connections between concepts. This approach ensures the SNOMED CT’s comprehensive medical knowledge structure, including its hierarchical and semantic relationships, is preserved within the graph database.

### MIMIC-IV Data Modeling

The MIMIC-IV data modeling approach focuses on creating a comprehensive graph structure that accurately represents the complex relationships in clinical data. The core entities are defined based on the primary tables in the MIMIC-IV database, with relationships designed to represent clinical hierarchies and preserve temporal associations between events.

The primary nodes represent essential clinical entities: Patient nodes use subject_id as their primary identifier, while Admission nodes preserve hadm_id along with temporal admission data. ICUStay nodes contain stay_id with corresponding admission and discharge times. Various clinical event nodes (e.g., Chart, Input, Output, Procedure) capture specific medical interventions and measurements. Item nodes serve as reference points for all clinical measurements and procedures, such as lab test results or medication dosages.

The relationships between nodes are designed to preserve the hierarchical (e.g., patients linked to admissions) and temporal nature (e.g., events ordered chronologically) of clinical data. Key relationships include HAS_ADMISSION connecting Patient to Admission nodes, HAS_ICU_STAY linking Admission to ICUStay nodes, and various event relationships connecting ICUStay nodes to their respective clinical events.

Clinical events are linked to their corresponding ICU stays rather than directly to patients, ensuring accurate temporal and contextual representation of each measurement or intervention. This systematic approach maintains data integrity while preserving critical clinical context. We devised this modeling approach to provide a foundation for comprehensive analysis of patient care pathways while maintaining efficient data access patterns for clinical decision support applications.

### SNOMED-CT Data Preprocessing

The processing of SNOMED CT data involved extracting and transforming the Release Format 2 (RF2) files from the July 2024 International Edition. While RF2 format includes various components, we specifically utilized three files: Concepts, Descriptions, and Relationships. Each component serves a distinct purpose in representing medical knowledge. Concepts form the foundational clinical entities, Descriptions provide human-readable terms, and Relationships define the semantic connections between concepts.

The integration process leveraged SNOMED-CT reference sets to map SNOMED-CT concepts to ICD-10-CM codes. This mapping process is crucial for maintaining semantic interoperability between clinical terminology systems, ensuring consistent interpretation of diagnostic concepts across different coding standards. The reference sets contain over 126,000 SNOMED CT source concepts mapped to ICD-10-CM targets, enabling comprehensive coverage of diagnostic concepts.

To transform the data into Neo4j’s graph structure, we preserved all original relationships from SNOMED CT without modification. The inherent hierarchical structure, including IS_A relationships and semantic relationships such as FINDING_SITE, CAUSATIVE_AGENT, and ASSOCIATED_WITH, was preserved exactly as defined in SNOMED CT’s original specification. This approach ensures the integrity of SNOMED-CT’s comprehensive medical knowledge representation while enabling efficient traversal and complex querying capabilities within the Neo4j.

### Integration

To integrate MIMIC-IV with SNOMED-CT, we created MAPS_TO relationships that link the icd_code property in MIMIC-IV’s diagnosis nodes to corresponding SNOMED-CT concepts. The mapping procedure involves normalizing ICD-10 codes, loading SNOMED-CT reference sets, and linking diagnostic codes to corresponding concepts.

The integration process begins by selecting diagnosis nodes in MIMIC-IV with ICD-10 codes. These codes are normalized to ensure consistent formatting, such as converting “I110” to “I11.0.” The normalized codes are then matched against SNOMED CT’s reference set to identify corresponding concepts. We utilized Neo4j’s MERGE operations to create and update SNOMED CT concept nodes. This process established MAPS_TO relationships between diagnosis nodes and their corresponding concepts.

This relationship structure enables advanced querying capabilities, such as exploring diagnostic patterns or tracing semantic connections between clinical events in MIMIC-IV and SNOMED-CT’s standardized medical terminology.

### Computational Constraints and Subset Analysis

Due to hardware limitations in this proof-of-concept study (MacBook with 16GB RAM), the complete MIMIC-IV dataset was processed in manageable subsets. The initial framework development utilized 1,504 patients to establish the graph structure and validate integration protocols. Subsequent clinical analyses were performed on relevant subsets extracted through specific inclusion criteria for each use case.

## Results

The integration of MIMIC-IV clinical data with SNOMED-CT resulted in a comprehensive graph database comprising 625,708 nodes, 2,189,093 relationships, and 114 properties across both datasets (Table 1). The primary labels include Patient, Admission, ICUStay, Diagnosis, and various clinical event types from MIMIC-IV, as well as Concept nodes from SNOMED-CT. Relationships such as HAS_ADMISSION and MAPS_TO preserve hierarchical and semantic connections between clinical entities. Figure 1 provides an example of a patient’s clinical trajectory with associated diagnoses and procedures.

**Table 1.**
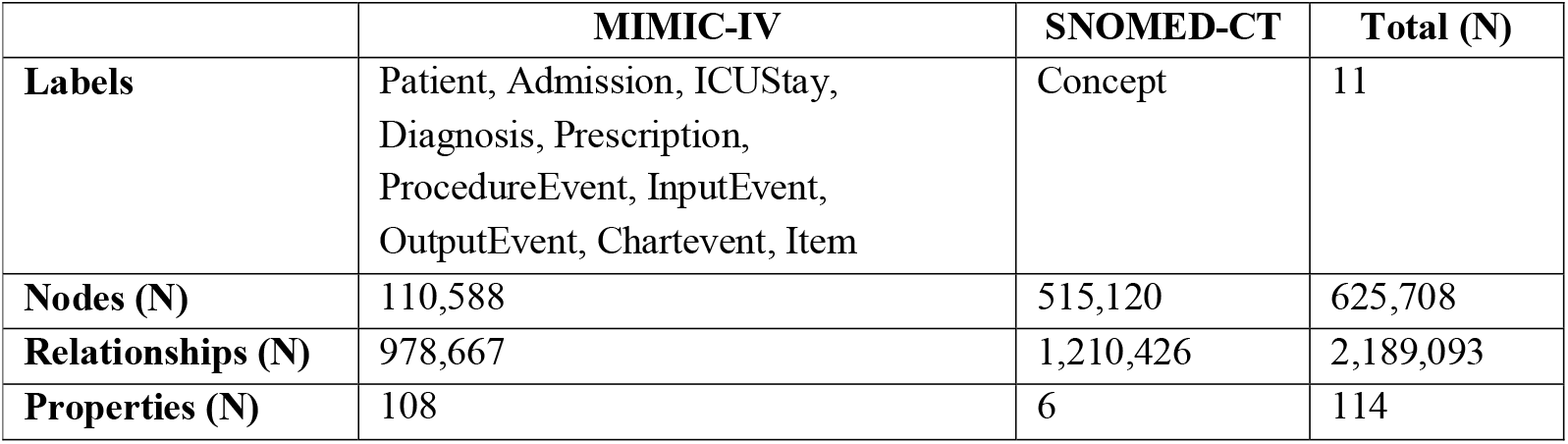
Distribution of Graph Database Components across MIMIC-IV and SNOMED-CT Integration.

**Figure 1.**
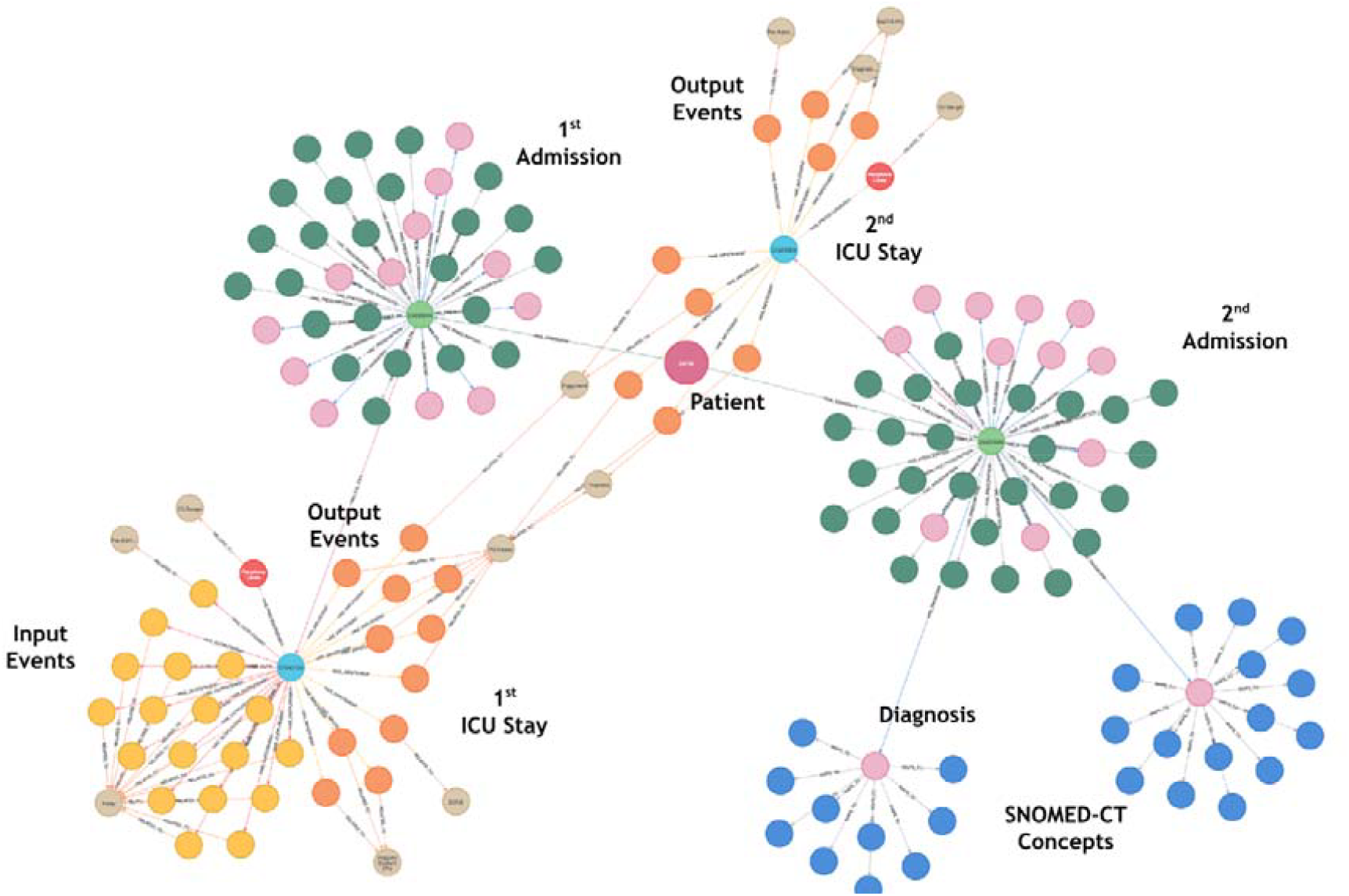
Sample Graph Visualization showing a patient’s clinical trajectory from admission through ICU stays with associated diagnoses and events, demonstrating temporal relationships and SNOMED-CT concept mappings

### Performance Test

To evaluate the technical advantages of the graph-based approach over traditional relational databases in handling complex healthcare data relationships, we conducted multiple performance tests comparing Neo4j and PostgreSQL across five query types. The evaluation encompassed three fundamental query types—Basic Retrieval, Path Analysis, and Pattern Matching—and two specialized clinical analyses designed to reflect common use cases in healthcare data analysis. Basic retrieval queries focused on identifying patient diagnoses for hyperlipidemia, while path analysis traced care trajectories from diagnosis to ICU procedures. Pattern matching queries evaluated patient diagnosis histories and ICU procedure patterns. The specialized analyses included evaluations of length of stay and disease correlations. The tests were performed with the following specifications:

*MacOS: Sequoia 15.3*

*Processor: Quad-Core Intel Core i5 @ 2.30GHz*

*Memory: 16GB 2133 MHz LPDDR3*

*Graphics: Intel Iris Plus Graphics 655, 1536BM*

PostgreSQL version 14.15 (installed via Homebrew) and Neo4j Community Edition version 5.24.0 were installed on this computer. Neo4j was the latest version available at the time of the study. The Neo4j Python driver version 5.27.0 was used for database connectivity, and Python 3.8 or higher was required to run the associated scripts. Neo4j was configured with an initial and maximum heap memory size of 3500MB and a page cache size of 1900MB.

The graph-based approach demonstrated consistently superior performance across all scenarios, with improvement factors ranging from 5.4x to 48.4x (Table 2). This substantial performance advantage is attributed to Neo4j’s native graph processing capabilities, which allow direct traversal of relationships without requiring expensive join operations. In contrast, PostgreSQL’s relies on multiple tables joins and complex aggregation operations, leading to significantly higher execution times as query complexity increases [25,26,27].

**Table 2.** Performance Comparison between Neo4j and PostgreSQL.

| Query Type | Neo4j (ms) | PostgreSQL (ms) | Improvement Factor |
| --- | --- | --- | --- |
| Basic Retrieval | 6 | 286 | 47.7x |
| Path Analysis | 9 | 290 | 32.2x |
| Pattern Matching | 26 | 1,258 | 48.4x |
| Length of Stay Analysis | 103 | 561 | 5.4x |
| Disease Correlation Analysis | 108 | 2,555 | 23.6x |

For instance, pattern matching queries in PostgreSQL required multiple join operations across patients, diagnosis, ICU stay, and procedure tables, resulting in execution times nearly 50 times slower than Neo4j’s direct relationship traversal approach. This dramatic improvement enables faster analysis of complex clinical patterns, supporting real-time decision-making and large-scale healthcare research.

### Clinical Applications

#### Temporal Analysis: Ventilator-Associated Pneumonia

The graph-based representation enabled analysis of temporal relationships between ventilator use and pneumonia in ICU settings. The analysis revealed that among 1,808 ventilated ICU stays, approximately 47.79% were associated with pneumonia, while among 2,252 non-ventilated ICU stays, approximately 47.16% were associated with pneumonia. It is important to note that these counts represent ICU stays or admissions rather than unique patients, as the dataset includes 1,504 unique patients, some of whom experienced multiple admissions or ICU stays.

This temporal pathway analysis was achieved through a query structure that traces patient pathways through admissions to ICU stays, identifies ventilator use through procedure events, and detects pneumonia diagnoses using SNOMED-CT concept mapping (conceptId: “13645005”). The graph structure’s ability to maintain temporal relationships facilitated precise calculations of pneumonia incidence rates across ICU stays, demonstrating the framework’s capability for monitoring ICU care quality and identifying potential risk factors.

#### Semantic Network Analysis: Hypertension

Our approach revealed complex clinical associations through comprehensive analysis of hypertension-related patterns. The query structure explored multiple levels of SNOMED CT relationships (IS_A, FINDING_SITE, ASSOCIATED_WITH) starting from hypertension diagnoses (ICD-10 code starting with ‘I10’). Through traversing up to three levels of relationships, we mapped anatomical correlations (e.g., systemic circulatory structures) and disease classification hierarchies (e.g., hypertensive disorders).

For instance, Figure 2 illustrates how hypertension connects to “Systemic circulatory system structure” through FINDING_SITE relationships and broader cardiovascular diseases like “Hypertensive disorder, systemic arterial” through IS_A hierarchies. This semantic network analysis demonstrated the framework’s capability to uncover rich clinical insights through standardized medical terminology, providing a foundation for identifying potential comorbidities or complications associated with hypertension, such as systemic arterial damage or cardiovascular comorbidities.

**Figure 2.**
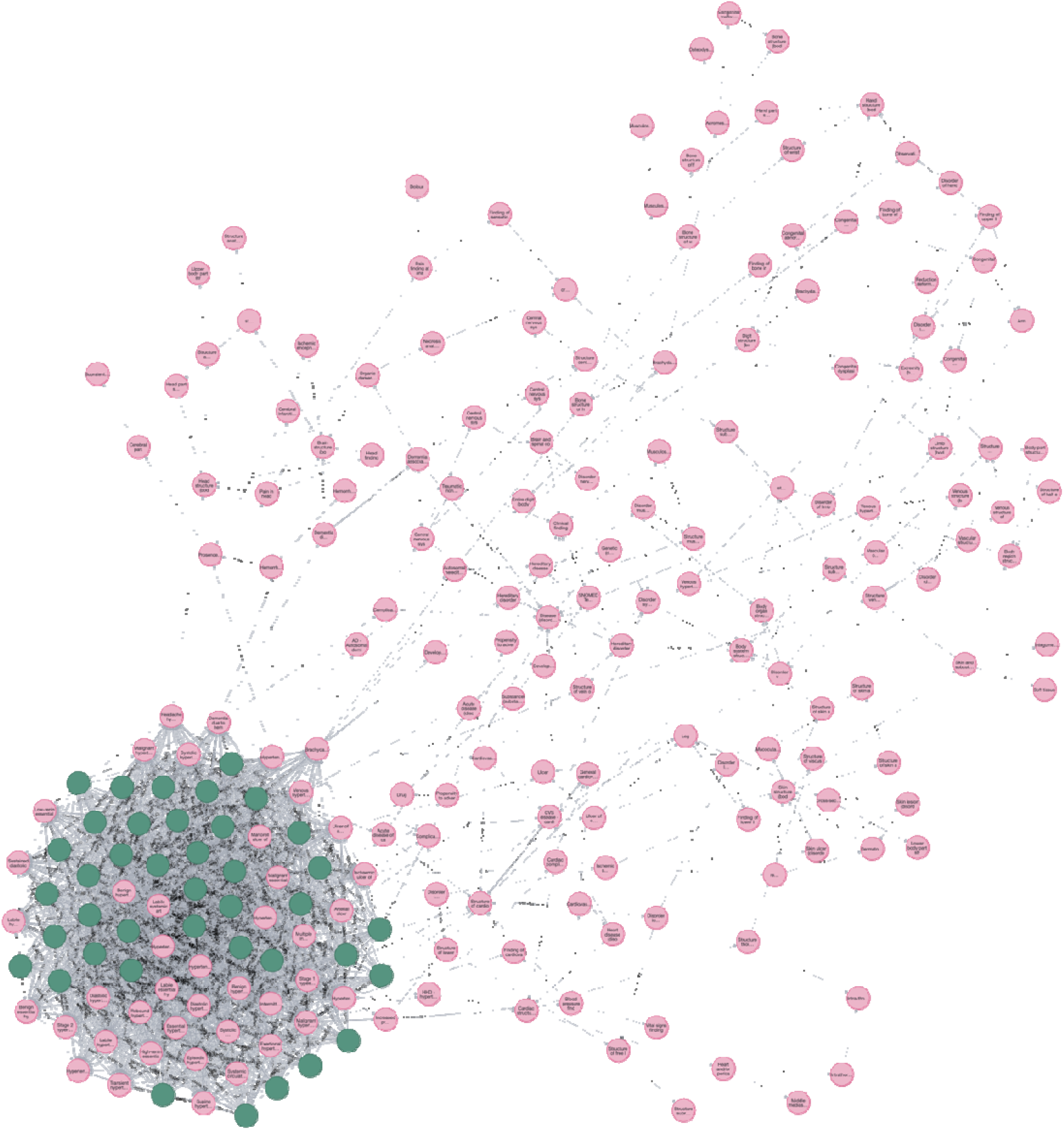
Semantic Network Analysis of Hypertension showing three-level relationship traversal through SNOMED-CT concepts (IS_A in blue, FINDING_SITE in green, ASSOCIATED_WITH in red) starting from ICD-10 code ‘I10’

In addition, leveraging this graph-based representation as a Retrieval-Augmented Generation (RAG) knowledge base for Clinical Decision Support Systems (CDSS) could enable more dynamic and context-aware reasoning by integrating semantic relationships into real-time decision-making processes. A CDSS could use the graph to answer questions like “What are common complications associated with hypertension?” or “Which anatomical sites are most affected by hypertensive disorders?” by dynamically retrieving relevant nodes and relationships.

#### Multi-dimensional Analysis: Drug Combinations

The graph database enabled analysis of drug combination outcomes through sophisticated query structures that identify concurrent medication administration patterns. Using ‘EXISTS’ clauses to ensure temporal alignment, we analyzed vancomycin-meropenem combination therapy administered concurrently within the same ICU stay.

The analysis was limited by small sample size (n=7), precluding statistical significance testing. Among these patients, 4 out of 7 (57.14%) survived to discharge. While this preliminary finding suggests potential efficacy, larger studies are needed to establish definitive treatment outcomes. This analysis serves as a proof-of-concept for the framework’s capability to identify concurrent medication patterns and investigate complex treatment relationships while maintaining temporal and contextual precision, rather than providing clinical recommendations.

#### Quality Measure Analysis: Real-World Performance Monitoring in U.S. Healthcare

Medicare Part D quality measures present significant challenges in the U.S. healthcare system, particularly for Medicare Advantage plans. This framework demonstrates practical applications in addressing these challenges through comprehensive analysis of critical quality measures that impact both patient safety and healthcare outcomes. This analysis is particularly timely as only 40% of plans achieved 4 stars or above in 2025, marking the lowest level in five years.

We focused on two key Part D measures: Statin Use in Persons with Diabetes (SUPD) and Concurrent Use of Opioids and Benzodiazepines (COB). For SUPD analysis, which showed a low average rating of 2.8 in 2025, we utilized Neo4j graph queries to examine statin prescriptions among diabetic patients aged 40-75 years. Our analysis revealed detailed prescription patterns through ICD code mapping (including codes 250.xx for diabetes) and medication tracking (Table 3). The results showed that among eligible diabetic patients, a significant portion lacked statin prescriptions despite clinical guidelines (Table 4). The most prescribed statins were Atorvastatin and Pravastatin, with varying prescription frequencies across different patient demographics.

**Table 3.** Distribution of Diabetes-Related ICD Codes.

| ICD Code Pattern | Description | Frequency |
| --- | --- | --- |
| 250 | Type 2 diabetes without complications | 78.30% |
| 250.40-250.43 | Diabetes with renal manifestations | 12.40% |
| 250.60-250.63 | Diabetes with neurological manifestations | 5.80% |
| 250.80-250.83 | Diabetes with other specified manifestations | 3.50% |

**Table 4.** Analysis of Statin Prescriptions in Diabetic Patients (40-75 years) *Note: Analysis performed on subset of 152 patients meeting diabetes criteria (ICD codes 250.xx) and age requirements from the total framework population of 1,504 patients.

| Characteristic | Value |
| --- | --- |
| Total Patients Analyzed | 152 |
| Statin Prescribed | 5 (3.3%) |
| No Statin | 147 (96.7%) |
| Most Common Statins | Atorvastatin, Pravastatin |
| Medicare Beneficiaries | 89 (58.6%) |
| Age Range | 40-75 years |

The COB measure analysis addressed the critical issue of potentially dangerous drug interactions, focusing on concurrent prescriptions exceeding 30 days. The framework analyzed temporal overlap of opioids (morphine, oxycodone, hydrocodone, fentanyl) and benzodiazepines (lorazepam, diazepam, clonazepam). Due to limitations in prescription date and supply duration data, we used hospital admission and discharge dates as a proxy for medication exposure periods. The analysis identified specific cases of concern, including a 54-year-old female Medicare beneficiary with a 36-day concurrent exposure period (from admission to discharge) and a 69-year-old male Medicare beneficiary with a 31-day overlap, demonstrating the framework’s capability to identify high-risk prescription patterns. While this approach using admission periods provides valuable initial insights for proof-of-concept, future implementations with detailed prescription timing data would enable more precise overlap analysis.

## Discussion

The integration of clinical data with standardized medical terminology in a graph structure enables systematic analysis of clinical relationships and treatment patterns. The framework demonstrates the capability to efficiently analyze complex clinical scenarios, from identifying temporal relationships in medication administrations to tracking patient outcomes across multiple encounters. Performance analysis reveals substantial improvements over traditional relational databases, with Neo4j showing 5.4x to 48.4x faster query execution across various clinical scenarios. This performance advantage is particularly significant for complex queries involving multiple joins and temporal relationships, which are common in clinical research. The dramatic improvement in query performance, driven by Neo4j’s graph-native architecture and relationship-centric data processing [25,27], enables real-time analysis of clinical patterns, potentially supporting rapid clinical decision-making in practice.

Furthermore, the graph-based approach offers inherent scalability, allowing for seamless integration of additional medical terminologies and clinical data types without restructuring the existing framework. The way we integrated SNOMED-CT with MIMIC-IV demonstrates this flexibility, as shown through our successful implementation of three diverse clinical use cases. The ventilator-associated pneumonia monitoring system leverages temporal relationship analysis to track clinical outcomes, demonstrating potential for real-time patient monitoring applications. The semantic network analysis of hypertension reveals complex disease relationships through SNOMED CT’s hierarchical structure, providing a foundation for knowledge-based clinical reasoning systems. The survival analysis of combined drug therapy showcases the framework’s ability to identify potential therapeutic patterns, suggesting applications in medication decision support.

As a proof of concept for actual issues in U.S. healthcare, we analyzed the Medicare Part D measures with specific focus on SUPD and COB metrics, which directly impact healthcare organizations’ Star Ratings and financial performance. The graph database framework demonstrated the ability to proactively identify target patient populations through simple database queries. For SUPD analysis, we identified diabetic patients aged 40-75 years without statin prescriptions from a complex dataset containing various diagnosis codes and medication records. Similarly, for COB measure, we successfully tracked concurrent prescriptions of opioids and benzodiazepines exceeding 30 days through temporal relationship analysis.

The clinical significance of this analysis extends beyond mere quality metrics. The graph-based framework demonstrated capability in supporting real-time quality measure monitoring while enabling proactive intervention in high-risk cases. The ability to efficiently query and analyze complex healthcare data relationships shows the practical value of the study’s approach in addressing real-world healthcare challenges. This provides healthcare organizations with tools for better coordination between providers and insurers, ultimately supporting the transition toward more effective, patient-centered care delivery while improving Medicare Star Ratings performance. This implementation proves that our graph database framework can effectively support proactive quality measure monitoring through straightforward database queries, offering a practical solution for healthcare organizations struggling with Medicare Part D performance metrics.

While the implementation demonstrates the framework’s effectiveness using MIMIC-IV, the methodology can be extended to various clinical databases and real-world healthcare systems. The MIMIC ecosystem offers diverse clinical data types through its specialized modules, demonstrating the potential for comprehensive clinical data integration [38,39,40,41,42]. While these extensive datasets exist within the MIMIC ecosystem, our current implementation focuses specifically on the core MIMIC-IV clinical database containing patient demographics, diagnoses, procedures, and ICU data.

The increasing availability of specialized clinical databases such as VitalDB and INSPIRE [36,37] further demonstrates the growing emphasis on incorporating real-world data in AI-powered clinical research settings. These databases capture different aspects of patient care, from high-resolution physiological measurements to comprehensive perioperative data, suggesting opportunities for enhanced clinical pattern analysis in real-world healthcare environments. Our framework’s architecture provides a methodological foundation for healthcare institutions to integrate their diverse data sources while supporting dynamic clinical decision-making processes, enabling sophisticated patient-centric analysis through graph-based representations while maintaining essential security and accessibility requirements for clinical practice.

The framework’s flexible architecture enables integration with other medical ontologies and standardized coding systems such as LOINC, RxNorm, or UMLS, supporting broader semantic interoperability across healthcare domains. The recent standardization of Graph Query Language (GQL) as ISO/IEC 39075:2024 marks a significant milestone for graph database technologies [34], similar to how SQL standardized relational database operations. This development suggests that graph databases are transitioning from experimental to industry-standard status, potentially accelerating their adoption in healthcare informatics. The standardization enables database-agnostic query development and reduces vendor lock-in concerns [35], which could facilitate broader implementation of graph-based clinical data analysis frameworks.

This proof-of-concept study has several important limitations. First, computational constraints necessitated analysis of patient subsets rather than the complete MIMIC-IV dataset, potentially limiting generalizability. Second, the temporal analysis relied on admission/discharge dates as proxies for medication timing, which may not reflect precise prescription periods. Third, small sample sizes in certain analyses (e.g., drug combination study, n=7) precluded statistical significance testing. Future implementations should address these limitations through enhanced computational resources and more granular temporal data.

Future research directions should extend this foundation in several key areas. First, semantic relationship validation through clinical expert verification will ensure accurate knowledge representation. Second, developing standardized mapping protocols will facilitate consistent transformation of clinical data into graph structures. Finally, evaluating compatibility with additional medical terminologies while maintaining semantic consistency will expand the framework’s utility across different healthcare domains. This methodological approach enables sophisticated analysis of clinical data while considering computational resources and data access limitations, providing a robust foundation for next-generation clinical decision support systems.

## Conclusion

This study presents a comprehensive technical framework for integrating clinical data with standardized medical terminology through graph database technology. Our proof-of-concept successfully demonstrates the feasibility of transforming complex relational clinical data into a semantically enriched graph structure, preserving both temporal relationships and clinical context. The framework’s effectiveness was validated through three distinct clinical applications, showcasing its potential for supporting clinical research and decision-making processes.

While technical challenges in terminology mapping and data structure transformation need to be addressed, this approach provides a promising foundation for advanced clinical data analysis and knowledge discovery. The framework’s ability to maintain semantic relationships while preserving temporal context opens new possibilities for clinical pattern analysis and hypothesis generation. Furthermore, its inherent scalability allows for seamless integration of additional medical terminologies and clinical data types without requiring significant restructuring, making it well-suited for handling larger datasets or expanding use cases in diverse healthcare settings.

The demonstrated performance improvements in query processing, combined with the flexibility of graph-based representation, suggest practical applicability in clinical research settings. Future development should focus on automated validation methods, integration with additional medical terminologies, and implementation of real-time clinical decision support features, ultimately moving towards more sophisticated and comprehensive clinical knowledge representation systems that can be readily integrated into existing healthcare infrastructures.

## Data Availability

The MIMIC-IV dataset used in this study is publicly available via PhysioNet (https://physionet.org/content/mimiciv/3.1/) upon completion of the required CITI training course. SNOMED-CT data is accessible through a licensing agreement with SNOMED International (https://www.snomed.org).

